# Genetic Transformer: An Innovative Large Language Model Driven Approach for Rapid and Accurate Identification of Causative Variants in Rare Genetic Diseases

**DOI:** 10.1101/2024.07.18.24310666

**Authors:** Lungang Liang, Yulan Chen, Taifu Wang, Dan Jiang, Jishuo Jin, Yanmeng Pang, Qin Na, Qiang Liu, Xiaosen Jiang, Wentao Dai, Meifang Tang, Yutao Du, Dirong Peng, Xin Jin, Lijian Zhao

**Author notes:** ***Corresponding author:*** Xin Jin, Lijian Zhao, ***E-mail:***. These authors contributed equally to this work (Lungang Liang, Yulan Chen, Taifu Wang, Dan Jiang).

## Abstract

**Background:** Identifying causative variants is crucial for the diagnosis of rare genetic diseases. Over the past two decades, the application of genome sequencing technologies in the field has significantly improved diagnostic outcomes. However, the complexity of data analysis and interpretation continues to limit the efficiency and accuracy of these applications. Various genotype and phenotype-driven filtering and prioritization strategies are used to generate a candidate list of variants for expert curation, with the final report variants determined through knowledge-intensive and labor-intensive expert review. Despite these efforts, the current methods fall short of meeting the growing demand for accurate and efficient diagnosis of rare disease. Recent developments in large language models (LLMs) suggest that LLMs possess the potential to augment or even supplant human labor in this context.

**Methods:** In this study, we have developed Genetic Transformer (GeneT), an innovative large language model (LLM) driven approach to accelerate identification of candidate causative variants for rare genetic disease. A comprehensive evaluation was conducted between the fine-tuned large language models and four phenotype-driven methods, including Xrare, Exomiser, PhenIX and PHIVE, alongside six pre-trained LLMs (Qwen1.5-0.5B, Qwen1.5-1.8B, Qwen1.5-4B, Mistral-7B, Meta-Llama-3-8B, Meta-Llama-3-70B). This evaluation focused on performance and hallucinations.

**Results:** Genetic Transformer (GeneT) as an innovative LLM-driven approach demonstrated outstanding performance on identification of candidate causative variants, identified the average number of candidate causative variants reduced from an average of 418 to 8, achieving recall rate of 99% in synthetic datasets. Application in real-world clinical setting demonstrated the potential for a 20-fold increase in processing speed, reducing the time required to analyze each sample from approximately 60 minutes to around 3 minutes. Concurrently, the recall rate has improved from 94.36% to 97.85%. An online analysis platform iGeneT was developed to integrate GeneT into the workflow of rare genetic disease analysis.

**Conclusion:** Our study represents the inaugural application of fine-tuned LLMs for identifying candidate causative variants, introducing GeneT as an innovative LLM-driven approach, demonstrating its superiority in both simulated data and real-world clinical setting. The study is unique in that it represents a paradigm shift in addressing the complexity of variant filtering and prioritization of whole exome or genome sequencing data, effectively resolving the challenge akin to finding a needle in a haystack.

## Introduction

Rare genetic diseases are hereditary, most are chronic and many result in early death, predominantly consist of Mendelian disorders. Currently, approximately 10,000 Mendelian diseases have been identified[1], among which the number of rare genetic diseases to be 6,000-8,000[2, 3]. A conservative, evidence-based estimate for the population prevalence of rare genetic diseases of 3.5–5.9%, which equates to 263–446 million persons affected globally at any point in time[3]. Individuals afflicted with a rare genetic disease are likely to possess causative variants that are both rare and deleterious. Understanding how genomic alterations result in different disease-related phenotypes is fundamental to diagnosis of rare genetic diseases[4].

The advent of high-throughput sequencing technologies over the past two decades has significantly reduced the difficulty in obtaining genotype information, thereby enhancing the discovery of genes responsible for rare genetic diseases and improving diagnostic rates. This advancement, nevertheless, introduces the complexity of sifting through a vast array of variants to identify the few that are pathogenic, particularly with whole exome sequencing (WES) and whole genome sequencing (WGS)[5].

Filtering and prioritization strategies aim to balance between maximizing sensitivity and minimizing the number of candidate variants, encompassing both genotype-driven and phenotype-driven analyses[6]. Genotype-driven analysis typically results in several hundred SNV/INDEL variants remaining for consideration[7]. Phenotype-driven analysis often serves as a complement to genotype-driven analysis, further refining the prioritization of remaining variants by integrating phenotypic information, with the aim of elevating genes or variants related to the patient’s phenotype for priority review by experts. There are phenotype-driven methods have been developed to prioritize genes and variants, including VAAST[8], Phevor[9], Phen-Gen[10], PhenIX[11], Exomiser[9], Phenolyzer[12], Genomiser[13], Xrare[14], LIRICAL[15], AMELIE[16], GEM[17], MOON[18], Emedgene[19], and AI-MARRVEL[20], etc. Those methods rely on three elements: phenotypic ontologies, genotype-phenotype databases, and algorithms for semantic similarity based on ontologies. Calculating semantic similarity based on phenotypes obtained during medical examinations and corresponding ontology entries is challenging. Phenotypes are inherently multimodal, and phenotypic ontologies, primarily composed of textual entries, may lack expressive power to accurately record individual phenotypes. Additionally, phenotypic descriptions are often “hasty or imprecise” in real-world settings[21], making it difficult to translate them into standardized phenotypic ontology entries. Moreover, phenotypic overlap between diseases complicates the situation further. As a result, phenotype-driven approaches show less than optimal effectiveness in practice.

Large Language Models (LLMs) have been developed through self-supervised learning on extensive text corpora, which equips them with the ability to discern semantic relationships between concepts without reliance on phenotype ontologies. There have been reports on the performance of LLM in the context of gene prioritization[22, 23], shown that LLMs can be used to provide diagnostic support that helps in identifying plausible candidate genes. Although these studies have primarily tested the performance of basic models in gene-level prioritization, it is theoretically plausible that fine-tuned LLMs could achieve superior performance. However, research pertaining to this assertion remains unreported.

We designated Genetic Transformer (GeneT) as an innovative LLM-driven approach for the filtering and prioritization of candidate causative variants. This study investigates the potential of GeneT for candidate causative variant identification (**Figure 1**). We systematically evaluate the ability and accuracy of identifying candidate causative variants after fine-tuning various LLMs across gradients of different training data volumes. Based on the optimal fine-tuned model, we constructed Genetic Transformer by integrating training sets from diverse data sources. We provide GeneT for interpretation experts in real-world clinical setting to assist them in identification of candidate causative variants. An online analysis platform iGeneT was developed to integrate GeneT into the workflow of rare genetic disease data analysis.

**Figure 1:**
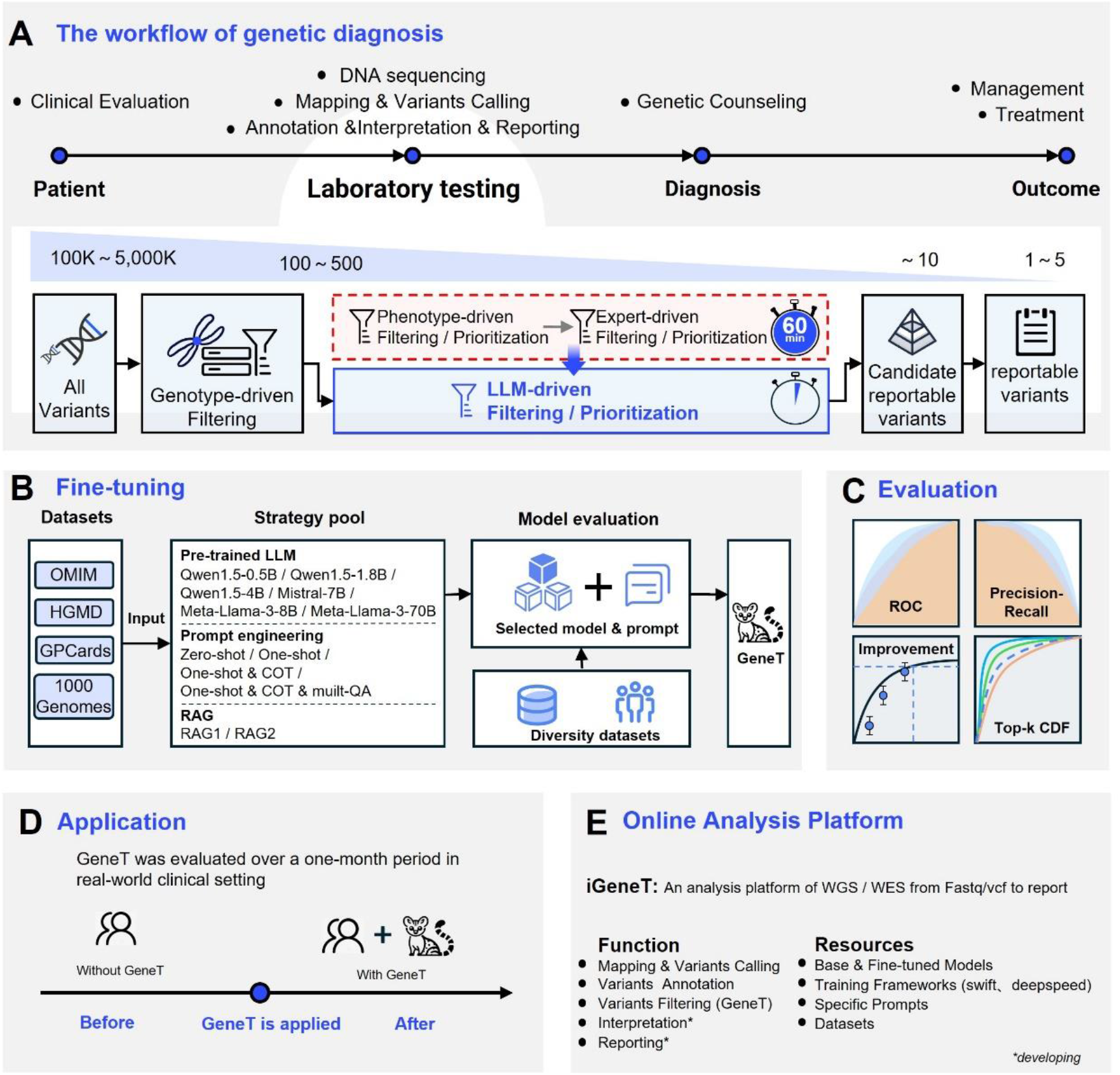
Study overview. **A)** The workflow of clinical genome sequencing applied to the diagnosis of rare genetic diseases. The newly developed LLM-driven filtering or prioritization methods hold promise in supplanting conventional approaches reliant on phenotype-driven and expert-driven methodologies. **B)** The fine-tuning process and optimization strategies of GeneT. **C)** The evaluation methods used; **D)** Application in real-world clinical setting. **E)** An interactive analysis platform incorporating GeneT, iGeneT.

## Result

### Fine-tuned LLMs enable accurate identification of disease-causing variations

We employed a training dataset TIO-1 and a testing dataset TIO-2, comprising synthetic cases from HGMD, OMIM, GPCards and 1000 Genomes Project, for fine-tuning and evaluating the LLMs (detail see methods). This fine-tuning process aimed to enhance the model’s proficiency in identifying candidate causative variants. Fine-tuning was performed on six pre-trained open source LLMs, including Qwen1.5-0.5B, Qwen1.5-1.8B, Qwen1.5-4B, Mistral-7B, Meta-Llama-3-8B, and Meta-Llama-3-70B. The training dataset, consisting of 12 gradients, ranges from 40 to 20,000 cases for the analysis of the scaling law of those models (**Table 1**). Figure 2 depicts the F1 scores of each model across varying training dataset sizes. When the training dataset size is 0, i.e., when the base model is used without fine-tuning, the F1 scores of all models are 0. These results are unable to effectively discern the pathogenicity of the variations, and within the scope of our study, they are consistently characterized as hallucinations. As the dataset size exceeds 800, the performance of those models stabilizes, with subsequent enhancements becoming insignificant, yielding F1 scores and AUC exceeding 0.9 (**Figure 2 A; Supplementary Table S1**). It is noteworthy that despite their smaller parameter size, both Qwen1.5-0.5B and Qwen1.5-1.8B exhibit a F1-score comparable to, or even superior to, that of other large-parameter models.

**Table 1:** F1 score of six pre-trained LLMs across varying sizes of training datasets.

| Training dataset size | Qwen1.5-0.5B | Qwen1.5-1.8B | Qwen1.5-4B | Mistral-7B | Meta-Llama-3-8B | Meta-Llama-3-70B |
| --- | --- | --- | --- | --- | --- | --- |
| 0 | 0.0000 | 0.0000 | 0.0000 | 0.0000 | 0.0000 | 0.0000 |
| 20 | 0.0000 | 0.0000 | 0.0000 | 0.0000 | 0.0000 | 0.0000 |
| 40 | 0.6667 | 0.0060 | 0.1022 | 0.6667 | 0.0000 | 0.0000 |
| 80 | 0.7904 | 0.8629 | 0.8885 | 0.6667 | 0.0526 | 0.0000 |
| 100 | 0.5690 | 0.8698 | 0.8870 | 0.6667 | 0.0479 | 0.0000 |
| 200 | 0.8876 | 0.9406 | 0.9323 | 0.6667 | 0.8655 | 0.0000 |
| 400 | 0.9290 | 0.9394 | 0.9480 | 0.8335 | 0.8625 | 0.8557 |
| 800 | 0.9426 | 0.9468 | 0.9520 | 0.9136 | 0.9470 | 0.9246 |
| 1000 | 0.9481 | 0.9586 | 0.9638 | 0.9272 | 0.9553 | 0.9348 |
| 2000 | 0.9589 | 0.9541 | 0.9729 | 0.9200 | 0.9467 | 0.9690 |
| 6000 | 0.9825 | 0.9831 | 0.9835 | 0.7930 | 0.9815 | 0.9811 |
| 10000 | 0.9795 | 0.9835 | 0.9875 | 0.9559 | 0.9771 | 0.9860 |
| 20000 | 0.9925 | 0.9925 | 0.9905 | 0.9075 | 0.9870 | 0.9890 |

**Figure 2:**
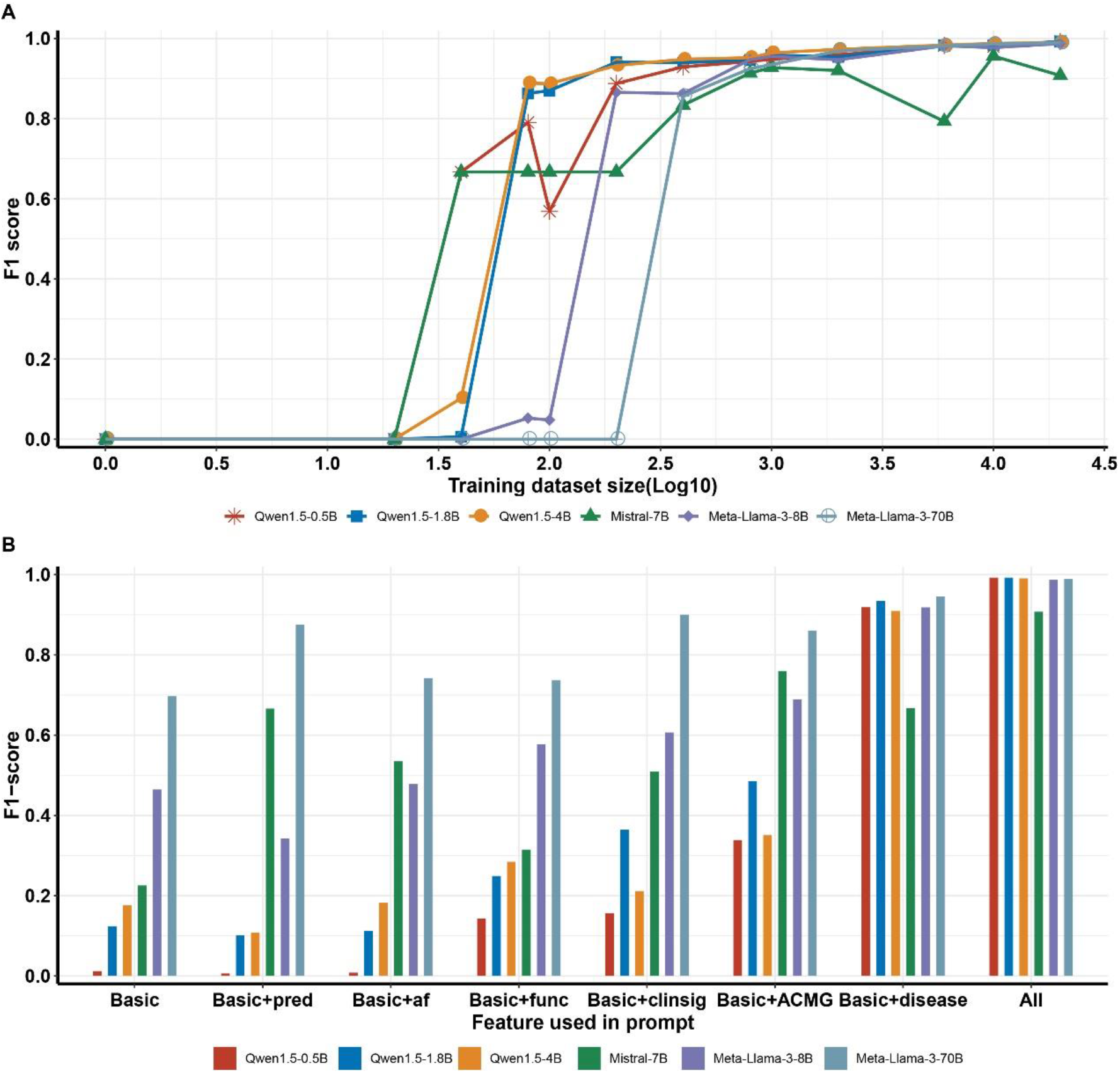
Fine-tuned LLMs enable accurate identification of disease-causing variations. **A)** Performance of fine-tuned LLMs across varying training dataset sizes. **B)** The F1 scores of the fine-tuned LLMs across different features.

Our model’s input prompts encompass a diverse array of variant features, including basic features (such as gene symbol, transcript, zygosity and HGVS name), functional data, ClinVar clinical significance, gene-associated disease information, population data, and ACMG interpretation information (**Supplementary Table S2**). To delve deeper into the learned feature importance of the model, we sequentially incorporated additional features into the inference of those fine-tuned models under the condition of variant basic information, to observe the changes in the F1 score. When only basic information is provided, we observed that models with a larger size of parameters perform better. For example, the model Qwen1.5-0.5B with a smaller size of parameters achieved an F1-score of only 0.0119, while Meta-Llama-3-70B reached 0.6969 (**Figure 2B; Supplementary Table S2**). This may be due to the fact that larger models possess a certain reservoir of pathogenic variant knowledge, which plays a positive role in determining pathogenic variants. Subsequently, the inclusion of various variant features led to an improvement in the F1 score, with the addition of gene-associated disease information producing the most significant enhancement. For instance, for Qwen1.5-0.5B, its F1 score increased from 0.0119 to 0.9188 (**Figure 2B**). Similar trends were observed in other models. This underscores the pivotal role of gene-associated disease information in the model’s assessment of pathogenicity.

### GeneT outperforms state-of-the-art methods on synthetic datasets

We then fine-tuned the six pre-trained LLMs, using the diversity training dataset TIO-3 (**see methods**). In GeneT, patient phenotypes are associated with variant information through prompt, and phenotypes are provided in various forms, such as disease names, free-text, and HPO/phenotype terms. We generated 200 synthetic whole-genome sequencing (WGS) samples for each of the three categories by leveraging data sourced from the HGMD, OMIM, and GPCards databases, alongside samples obtained from the 1000 Genomes Project (**see methods**). This methodology was employed to assess the model’s performance across diverse phenotypic data sets.

After the implementation of GeneT, the number of variants in the three test set samples decreased significantly, most notably for Qwen1.5-1.8B, from an average of 421, 414, and 419 to 9, 5, and 10, respectively. The Mistral-7B performed the least well, with more false-positive variants detected (**Figure 3 A, B, C**). Meanwhile, the fine-tuned Qwen1.5-1.8B achieved positive variant recalls of 0.98, 0.99 and 0.98 in the three test sets, respectively. Other fine-tuned LLMs exhibit comparable or marginally inferior performance **(Supplementary Tables S3-8)**. These results indicate that GeneT has the capacity to substantially decrease the number of candidate causative variants while upholding a high recall rate for causative variants.

**Figure 3:**
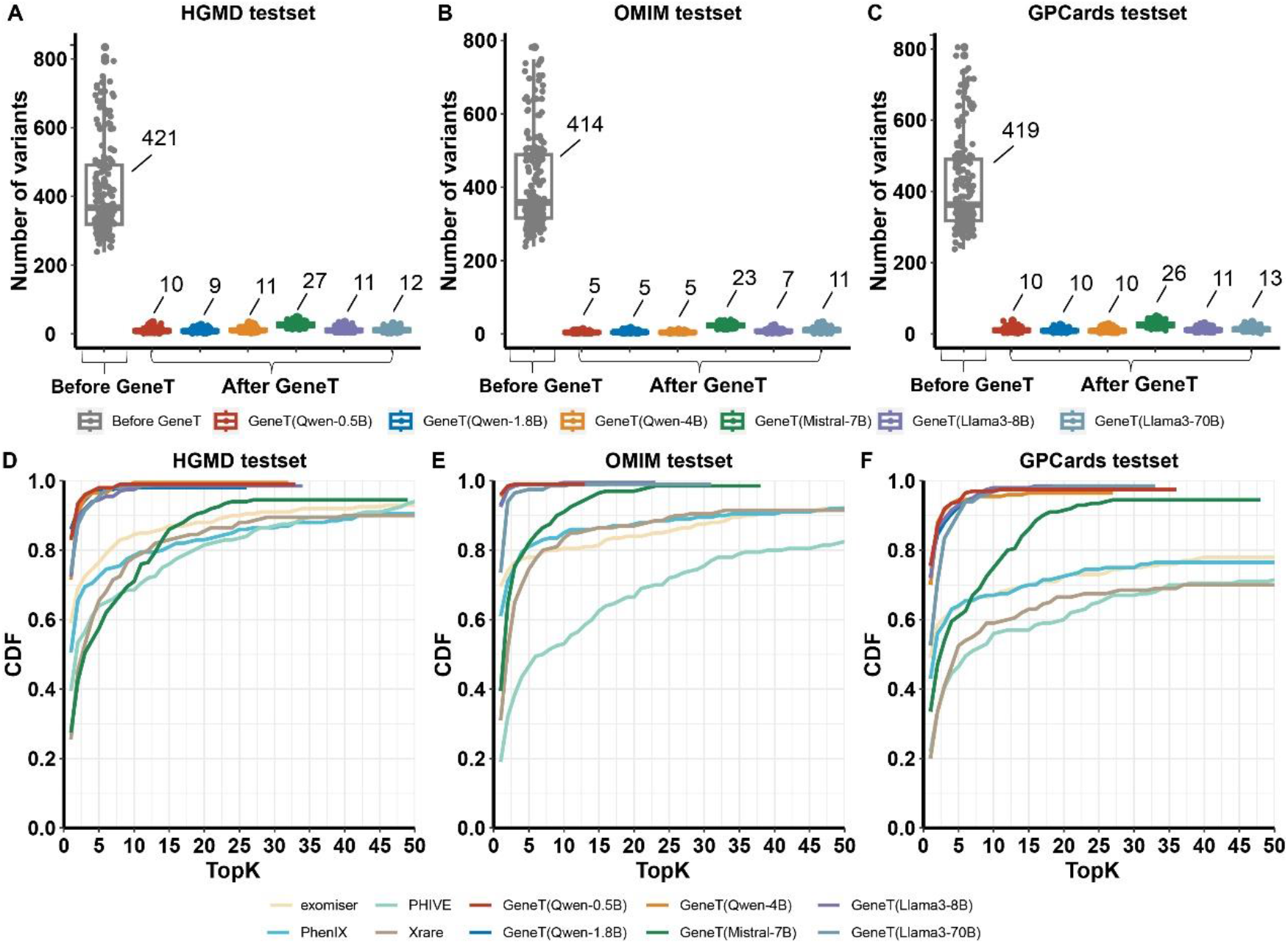
Performance evaluation on synthetic datasets. The number of candidate variants before and after the utilization of GeneT and TopK CDF plots on **AD)** the HGMD-1000Genomes dataset (HKG-I), **BE)** the OMIM-1000Genomes dataset (OKG-I), **CF)** the GPCards-1000Genomes dataset (GKG-I). CDF: Cumulative Distribution Function. The numerical values adjacent to each boxplot represent the mean number of candidate variations.

GeneT was then compared with existing state-of-the-art phenotype-driven variant prioritization methods on the synthetic samples for performance evaluation. We ranked the positive variants identified by GeneT using the predicted token probabilities to calculate the cumulative distribution of TopK recall performance. The results suggest that GeneT has demonstrated significantly superior performance compared to other phenotype-driven methods across the three test sets (**Figure 3D, E, F, Supplementary Tables S3-8**). Except for Mistral-7B, the performance of other fine-tuned LLMs is comparable, achieving high cumulative recall (>0.9) at lower TopK thresholds. Additionally, the token probabilities predicted by GeneT can be employed to prioritize all candidate variants within each sample, extending beyond the variants predicted as positive by GeneT. Upon prioritizing all variants based on these probability values, we noted that variants predicted as negative but actually positive were positioned at relatively higher ranks (**Supplementary Figure S1, Tables S3-8**). This indicates that GeneT can also serve as an alternative for prioritizing variant ranking tools.

### An application in real-world clinical setting confirms GeneT markedly boosts expert efficiency and precision in identifying candidate variants

We provided GeneT as a plugin in our interpretation system for interpretation experts in real-world clinical setting to assist them in identification candidate causative variants. The efficacy of the GeneT was then evaluated over a one-month period within the real-world clinical setting. In the realm of clinical genetic testing, the assurance of interpretation result reliability often involves the allocation of expert 1 and expert 2 for the initial screening of candidate variations and subsequent review of the preliminary screening outcomes. GeneT is principally employed to aid expert 1 in the process of screening for genetic variations. The results indicated a significant reduction in analysis time when utilizing the GeneT results while adhering to the standard operating procedure (SOP). The average analysis time decreased from 63.22 minutes to 44.11 minutes per sample (p=2.57e-13) **(Figure 4A**). Additionally, if GeneT results are used directly, there is potential for a 20-fold increase in speed, reducing the time required from approximately 60 minutes to around 3 minutes per sample. Furthermore, a marginal enhancement in the recall rate was observed (p=0.0846), rising from 94.36% to 97.40%. Concurrently, the variance in recall rates among interpretation experts decreased (**Figure 4B**). The outcomes of this prospective investigation validate the capacity of GeneT to augment interpretive efficiency and precision. These findings emphasize the significance of GeneT as a valuable component within the genetic analysis workflow.

**Figure 4.**
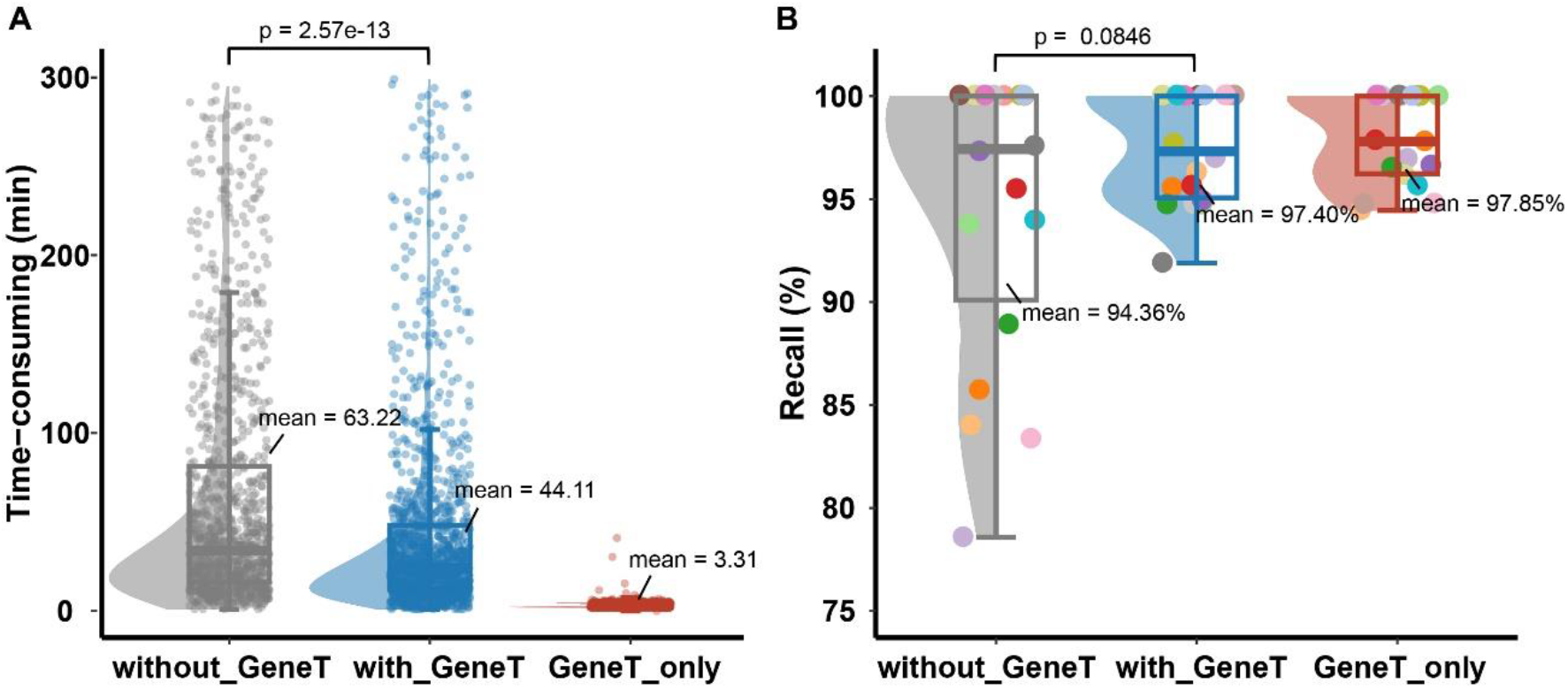
The effectiveness of GeneT in real-world clinical setting. **A)** The Time consumption and **B)** Recall rates for interpretation experts with and without the use of GeneT, alongside with GeneT_only recall performance. Each data point in the left graph corresponds to a sample, whereas the points in the right graph denote interpretation experts. Without_GeneT: Experts in interpretation refrain from utilizing GeneT and instead interpret based on their Standard Operating Procedures (SOP). Without_GeneT: Interpretation experts utilize GeneT as a point of reference while adhering to their original SOP. GeneT_only: Solely employs GeneT analysis, with the time denoting the duration for analyzing a single sample using GeneT.

### Hallucinations and reproducibility

Hallucination is a prevalent issue in LLMs, while reproducibility is a metric of significant importance in clinical applications. The assessment of hallucination and reproducibility is a crucial aspect of LLM utilization in clinical diagnostics. In this paper, we define the hallucination as outcomes incongruent with the anticipated outputs of “positive” or “negative”. We observed that the pre-training LLM exhibited significant hallucination issues before to fine-tuning, despite our constraints on the model’s output, which were notably improved after fine-tuning, particularly as the size of the fine-tuning dataset gradually increased (**Figure 5, Supplementary Table S1**). One of the primary reasons for this improvement lies in our transformation of the challenge of screening or ranking the entire pathogenic variation into an individual variation pathogenicity classification problem. Additionally, we enhanced the prompt by incorporating the “COT” directive, enabling the model to engage in step-by-step self-verification and improving its performance in multi-step reasoning tasks. The prompt also served to restrict the model’s output, allowing it to produce only “negative” or “positive” results. Furthermore, the integration of the RAG strategy, which links to external knowledge bases, bolstered the model’s ability to validate answers and to some extent, mitigated the generation of hallucinations.

**Figure 5.**
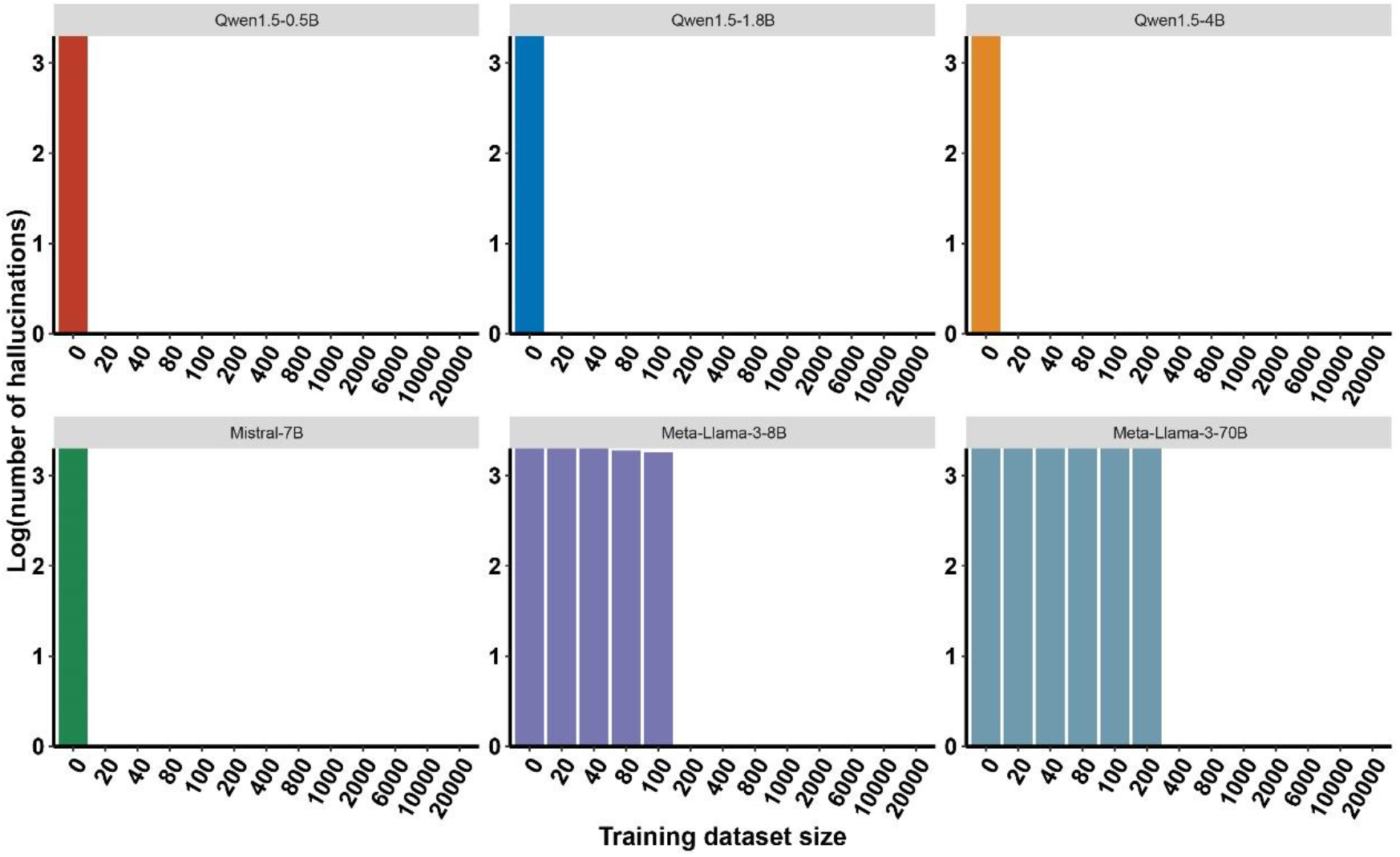
Hallucinations of six LLMs. The abscissa denotes the magnitude of the training dataset, while the ordinate signifies the occurrence of hallucinations. As the size of training data increases, the hallucinations of the model gradually dissipate.

During text generation, the LLM utilizes probability distributions to select the next word, thereby introducing a certain degree of randomness. To maximize the stability of LLM output, we set the temperature parameter of the softmax function in the LLM to 0. We utilized GeneT to perform inference analysis on a randomly chosen set of six genetic variants, executing a total of 10,000 replicate experiments for each to evaluate the variability in the predictive results. The results of these experiments exactly the same, indicate a high level of consistency and reproducibility in the identification of candidate variants by GeneT.

### iGeneT: an interactive analysis platform incorporating GeneT

We developed an interactive analysis platform iGeneT (http://igenet.genomics.cn), which integrates GeneT, enabling users to perform variants analysis using an online web interface. Users can perform analysis by submitting the sample’s clinical phenotype and VCF file.

## Discussion

We present GeneT as an innovative LLM-driven approach for the filtering and prioritization of candidate causative variants. Our results demonstrate an exceptional performance compared to existing state-of-the-art phenotype-driven variant prioritization methods, achieving the recall rates of 99% and 98% in synthetic datasets and real-world clinical setting, respectively. Additionally, the results indicate the potential for a 20-fold increase in speed, reducing processing time from approximately 60 minutes to around 3 minutes per sample.

In this study, GeneT supports phenotypes presented in various forms, such as disease names, free-text descriptions, and HPO/phenotype terms. In contrast to phenotype-driven methods, LLMs possess the capability to discern semantic relationships between concepts without relying on phenotype ontologies. Moreover, LLMs have the potential to leverage additional phenotypic information, including medical records, speech, images, and radiographs, which can not only facilitate the identification of candidate variants but also help prevent diagnostic delays [24].

This study concentrates on the primary findings at the single-variant loci level, including single nucleotide variants (SNVs) and insertions/deletions (InDels) within the protein-coding regions. GeneT is also discernible, albeit with slightly lower performance in other contexts currently. These contexts include (1) secondary findings, which are results that are not related to the indication for ordering the sequencing but that may nonetheless be of medical value or utility to the ordering physician and the patient. [25]; (2) other types of variations within coding regions, as well as those in other regions, such as copy number variations (CNVs), repeat expansions, and structural variations (SVs), which are anticipated to account for part of the remaining positive cases[26]; (3) compound loci situations, such as compound heterozygosity[27, 28], complex compound patterns[29, 30], and digenic inheritance[31, 32]. However, these scenarios were not specifically trained for or evaluated in the current study and will be the focus of future model enhancements.

Our study reveals that a model with 0.5 and 1.8 billion parameters demonstrates optimal performance, suggesting that smaller parameter size LLMs may be more adept than larger parameter size LLMs for executing binary classification tasks. The prevailing notion that “bigger is better”[33] has been a central theme in the recent evolution of LLMs. While our study also indicates that larger parameter size LLMs exhibit stronger overall capabilities (**Figure 2B**), they may not be ideally suited for applications requiring on-device processing capabilities, energy efficiency, a minimal memory footprint, and swift response times. These attributes are crucial for ensuring privacy, security, and the sustainable deployment of AI-driven solutions.

In conclusion, we present GeneT as an innovative LLM-driven approach for the rapid and accurate identification of causative variants in rare genetic diseases. This research represents a paradigm shift in the methodology for interpreting genetic disorders. By leveraging the extensive knowledge base of large language models (LLMs), their advanced semantic comprehension, and the capacity for comprehensive training and fine-tuning, we can integrate the analytical reasoning and specialized expertise of domain professionals. Beyond its application in the identification of candidate causative variants, the powerful potential of LLMs extends to assess the risk of genetic disorders based on phenotype, variant interpretation, and genetic counseling. This approach holds the promise to replace traditional algorithms and either augment or even supplant human labor in these contexts, thereby accelerating the diagnosis of rare genetic diseases.

## Material and methods

### Datasets

We have established multiple training and testing datasets for model fine-tuning and testing, constructed from data sources, including HGMD[34], OMIM[35], GPCards[36], and the 1000 Genomes Project[37]. All variants have been annotated by an in-house pipeline, integrating tools such as BCFanno[38] and SIGVAR[39]. To mitigate the influence of the training set on the testing set, it is ensured that the testing set comprises variants not present in the training set.

#### The HGMD-1000Genomes Dataset

Based on the HGMD Professional database release 2018.3, we selected DM-marked variants absent from the ClinVar database. Utilizing these variants, we have constructed two datasets (as detailed below), specifically designated for model fine-tuning and testing.

- **HKG-T**: 26826 cases, with phenotype and pathogenic variant from HGMD, non-pathogenic variants from 1000 Genomes.
- **HKG-I**: 200 synthetic WGS cases using phenotype and pathogenic variants randomly extracted from HGMD inserted into genome-wide VCF files released by the 1000 Genome Project. Through variant annotation and preliminary filtering analysis, nearly 400 candidate variants remained for each sample.

#### The OMIM-1000Genomes Dataset

Based on the archived OMIM updated on February 22, 2011(*https://ftp.ncbi.nih.gov/repository/OMIM/ARCHIVE)*, we extracted the descriptions in the “CLINICAL FEATURES” section from omim.txt.Z file as phenotypes and matched them with the variants on pathogenic genes in ClinVar. We have constructed two datasets (as shown below), designated for model fine-tuning, and TopK evaluation, respectively.

- **OKG-T**: 19770 cases, with phenotype and pathogenic variant from OMIM and ClinVar, non-pathogenic variants from 1000 Genomes.
- **OKG-I**: 200 synthetic WGS samples using phenotype and pathogenic variants from OMIM (manually curated post 2012) inserted into genome-wide VCF files released by the 1000 Genome Project. Through variant annotation and preliminary filtering analysis, nearly 400 candidate variants remained for each sample.

#### The GPCards-1000Genomes Dataset

Based on the records downloaded from GPCards, we documented a table file of sample phenotypes and genotypes, from which we extracted a subset for use in this study. We excluded variants recorded in ClinVar and, based on phenotype deduplication, randomly selected 200 variants to serve as the testing set, with the remainder constituting the training set. We have constructed two datasets (as shown below), designated for model fine-tuning, and TopK evaluation, respectively.

- **GKG-T**: 8748 cases, with phenotype and pathogenic variant from GPCards, non-pathogenic variants from 1000 Genomes.
- **GKG-I**: 200 synthetic WGS samples using phenotype and pathogenic variants from GPcards inserted into genome-wide VCF files released by the 1000 Genome Project. Through variant annotation and preliminary filtering analysis, nearly 400 candidate variants remained for each sample. The testing set has variants that are not found in the training set.

#### Three-in-One Datasets

The diversity of the training data is a crucial factor in enhancing model performance, as it enables the model to better understand and process various types of inputs, thereby achieving superior results in practical applications. To ensure the efficacy of model training, we constructed two comprehensive three-in-one datasets to enhance the diversity of the training set for fine-tuning the Large Language Model (LLM) (as shown below).

- **TIO-1**: 20000 cases, constructed from three distinct datasets in specific proportions: 25% OKG-T, 45% HKG-T, and 35% GKG-T. This dataset is utilized for gradient testing of the Large Language Model (LLM).
- **TIO-2:** 2000 cases, constructed from three distinct datasets in specific proportions: 25% OKG-T, 45% HKG-T, and 35% GKG-T. This dataset is utilized for gradient testing of the Large Language Model (LLM), all variants not present in the training set TIO-1.
- **TIO-3**: 55344 cases, integrating the OKG-T, HKG-T, and GKG-T datasets. It is employed for GeneT fine-tuning and TopK CDF assessment.

### Phenotype-driven methods

Four phenotype-driven tools were selected for comparison: PHIVE[40], PhenIX[11], Exomiser[9], and Xrare[14]. These have been chosen as benchmarks due to their demonstrated utility and the availability of comparative data, enabling a rigorous evaluation against other emerging tools in the field. This selection aims to provide a representative cross-section of the current state-of-the-art in phenotype-guided variant prioritization, facilitating a comprehensive analysis of their respective capabilities and shortcomings. The HPO terms input into the tools are uniformly extracted from the patient’s phenotype descriptions using PhenoTagger[41]. Xrare is executed using the xrare37-pub:2021 docker image. exomiser(v13.3.0) is executed using its default algorithm hiPhive, while phenIX and PHIVE are executed using the phenix and phive algorithms from the exomiser suite, respectively.

### Pre-trained LLM

We have chosen six diverse pre-trained Large Language Models (LLMs) for fine-tuning purposes, encompassing a spectrum of parameter sizes ranging from 0.5 billion to 70 billion. Specifically, the models selected are:

1. “Qwen1.5-0.5B”, “Qwen1.5-1.8B” and “Qwen1.5-4B”, which are pre-trained LLMs open-sourced by Alibaba[42]. These models are characterized by a relatively modest parameter size, offering a compact yet robust foundation for fine-tuning endeavors.
2. “Mistral-7B”, a large-scale model developed by the French startup Mistral AI. The model adopts advanced training techniques and a large amount of corpus data, providing powerful language processing capabilities and a wide range of applications. Whether it’s text generation, sentiment analysis, or question-answering systems, Mistral-7B can deliver outstanding performance.
3. “Meta-Llama-3-8B” and “Meta-Llama-3-70B,” representing the state-of-the-art in LLM technology and recently open-sourced by Meta[43]. These models are distinguished by their extensive parameterization and have demonstrated superior performance in various language tasks, positioning them as exemplary candidates for advanced fine-tuning applications.

The selection criteria were designed to ensure a diverse representation of LLMs across different parameter sizes and linguistic focuses, allowing for a nuanced investigation into the fine-tuning outcomes and potential optimizations across a wide array of applications.

### Prompt Engineering

We designed a structured prompts for LLM fine-tunning and inferencing, followed the GPT guidelines[44, 45] to design our prompts. We wrote clear instructions by following the suggested tactics (e.g., ask the model to adopt a role, use delimiters to clearly indicate distinct parts of the input, specify the output format).

**Supplementary Table S9** shows the prompts with which we experimented. Prompts P1 is zero-shot, P2 is few-shot, P3 constitutes a few-shot, chain-of-thought prompt instructing the LLM specifically on how to perform the variant identification. While P4 constitutes a one-shot, chain-of-thought prompt and RAG. In P1, we instruct the LLM to identify whether the variant is negative or positive. In P2, additional a few outputs example is provided. In P3, the LLM is prompted to identify whether the variant is negative or positive based on the chain of thinking. Lastly, P4, a few-shot, chain-of-thought prompt augmented by variant rich-annotated information.

To ascertain the optimal prompt for subsequent research and application endeavors, we conducted an evaluation of the performance of four distinct prompts using GPT-4, and identified P4 as the selected prompt.

### Fine-tuning LLM

In the process of fine-tuning Large Language Models (LLMs), we have systematically combined datasets, pre-trained LLMs to train a cohort of models for subsequent evaluation and application. This ensemble includes 78 models designated for gradient testing, encompassing 6 distinct LLMs, 13 variations across data gradients. Additionally, we developed another model that combine different training sets to enhance dataset diversity.

The training of these models was executed on a cluster comprising 2 high-performance GPU servers (8 CUDA-capable GPUs, namely NVIDIA A100 Tensor Core GPU with 80 GB graphic memory), leveraging the Swift training framework to facilitate the computationally intensive tasks associated with LLM fine-tuning. The full-parameter fine-tuning mode is used, with “sft_type” set to “full”, and training is set for 5 epochs.

### Evaluation

We implemented a training data size gradient analysis. This approach involves incrementally varying the amount of data used in the fine-tuning process to observe its impact on the performance. Specifically, the gradient analysis examines how the performance changes with different data sizes, offering insights into the robustness and scalability of the fine-tuned LLMs across a spectrum of data availability.

We conducted an assessment of the variant ranking performance utilizing the TopK cumulative distribution function (TopK CDF) as a primary metric. This statistical measure is widely recognized for its ability to consolidate the true positive rate and false positive rate across various threshold settings into a single value, thereby providing a comprehensive evaluation of the ranking efficacy.

### ETHICS APPROVAL AND CONSENT TO PARTICIPATE

The studies involving human participants were reviewed and approved by the Ethical Clearance the Institutional Review Board of BGI. Written informed consent for participation was not required for this study in accordance with the national legislation and the institutional requirements.

## Data Availability

All data produced in the present study are available upon reasonable request to the authors

https://ftp.ncbi.nlm.nih.gov/pub/clinvar

https://omim.org

https://ftp.ncbi.nih.gov/repository/OMIM/ARCHIVE/

https://www.hgmd.cf.ac.uk/ac/index.php

https://hgdownload.soe.ucsc.edu/gbdb/hg19/1000Genomes/

## Author Contributions

L.L., Y.C., T.W. and D.J. were involved in study conception and design, T.W., Y.C., L.L. performed data analysis and data interpretation, Y.C. and T.W. draft the manuscript, J.J., W.D., M.T., D.P. and Y.D. contributed to the application in real-world clinical setting, Y.P. and Q.N. contributed to the deployment and fine-tuning of LLM, X.J. contributed to visualization, Y.P. and Q.L. contributed to dataset preparation and iGeneT development, X.J., Y.D. and L.L. revised the manuscript, D.J. and L.Z. supervised the study. All authors read and approved the final manuscript.

