## Supplemental Figure S1 for "Genetic Transformer: An Innovative Large Language Model Driven Approach for Rapid and Accurate Identification of Causative Variants in Rare Genetic Diseases"

Supplementary Appendix

Table of Contents

Figure. S1: Performance evaluation on synthetic datasets.

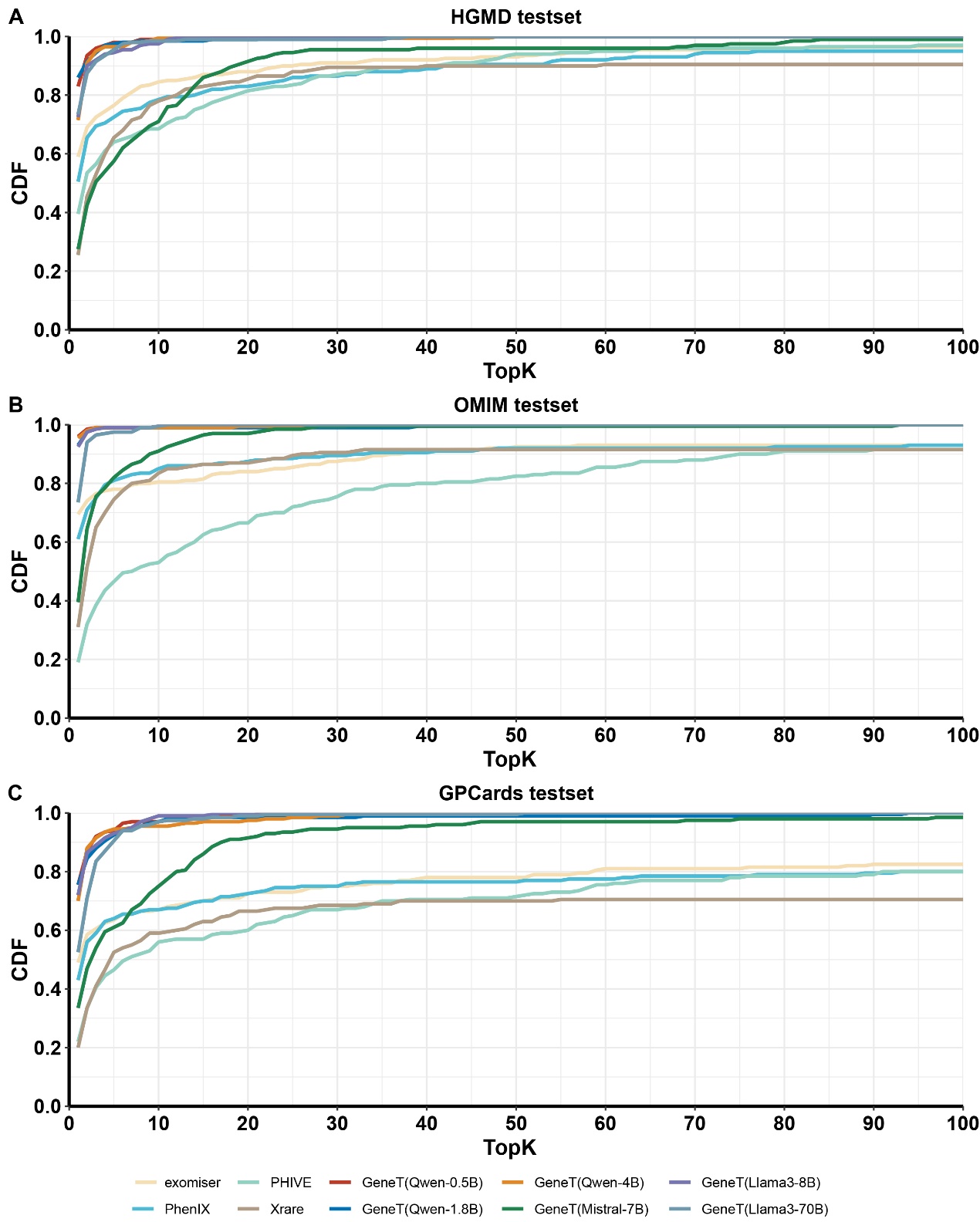

**Figure S1: Performance evaluation on synthetic datasets.** The TopK CDF plots on **A)** the HGMD-1000Genomes dataset (HKG-I), **B)** the OMIM-1000Genomes dataset (OKG-I), **C)** the GPCards-1000Genomes dataset (GKG-I). CDF: Cumulative Distribution Function.
